# C-reactive protein-based tuberculosis triage testing: a multi-country diagnostic accuracy study

**DOI:** 10.1101/2024.04.23.24305228

**Authors:** Brigitta Derendinger, Tessa K. Mochizuki, Danaida Marcelo, Deepa Shankar, Wilson Mangeni, Hanh Nguyen, Seda Yerikaya, William Worodria, Charles Yu, Nhung Viet Nguyen, Devasahayam Jesudas Christopher, Grant Theron, Patrick P.J. Phillips, Payam Nahid, Claudia M. Denkinger, Adithya Cattamanchi, Christina Yoon

## Abstract

**Rationale:** C-reactive protein (CRP)-based tuberculosis (TB) screening is recommended for people with HIV (PWH). However, its performance among people without HIV and in diverse settings is unknown.

**Objectives:** In a multi-country study, we aimed to determine whether CRP meets the minimum accuracy targets (sensitivity ≥90%, specificity ≥70%) for an effective TB triage test.

**Methods/Measurements:** Consecutive outpatient adults with cough ≥2 weeks from five TB endemic countries in Africa and Asia had baseline blood collected for point-of-care CRP testing and HIV and diabetes screening. Sputum samples were collected for Xpert MTB/RIF Ultra (Xpert) testing and culture. CRP sensitivity and specificity (5 mg/L cut-point) was determined in reference to sputum test results and compared by country, sex, and HIV and diabetes status. Variables affecting CRP performance were identified using a multivariate receiver operating characteristic (ROC) regression model.

**Results:** Among 2904 participants, of whom 613 (21%) had microbiologically-confirmed TB, CRP sensitivity was 84% (95% CI: 81-87%) and specificity was 61% (95% CI: 59-63%). CRP accuracy varied geographically, with higher sensitivity in African countries (≥91%) than Asian countries (64-82%). Sensitivity was higher among men than women (87% vs. 79%, difference +8%, 95% CI: 1-15%) and specificity was higher among people without HIV than PWH (64% vs. 45%, difference +19%, 95% CI: 13-25%). ROC regression identified country and measures of TB disease severity as predictors of CRP performance.

**Conclusions:** Overall, CRP did not achieve the minimum accuracy targets and its performance varied by setting and in some sub-groups, likely reflecting population differences in mycobacterial load.

## Introduction

Each year, one-third (4 million) of all individuals with active tuberculosis (TB) are not diagnosed or reported, leading to unfavorable outcomes for people with TB and ongoing TB transmission within communities.^1^ New approaches are urgently needed to reach the “missing 4 million” and reduce TB incidence worldwide. Triage testing can help prioritize higher risk individuals for confirmatory testing and an effective non-sputum triage test is considered among the highest priority needs for TB diagnostics.^2^ Modeling studies have shown that an effective TB triage test – defined as a simple-to-perform, point-of-care, low-cost test with a minimum sensitivity and specificity of 90% and 70%, respectively – could increase TB case detection while reducing unnecessary confirmatory testing by 40-60%.^3^

TB triage testing options are limited. Chest x-ray (CXR) has high accuracy but requires expensive infrastructure, has high per test costs, and requires either trained personnel or computer-aided detection algorithms for interpretation.^4–6^ No other tests meet the accuracy targets and operational characteristics of an effective TB triage test.

C-reactive protein (CRP), a non-specific marker of inflammation, was recently recommended by World Health Organization (WHO) as a TB screening tool to facilitate intensified case finding among people living with HIV (PWH) based on multiple diagnostic accuracy studies and an individual patient meta-analysis.^7^ However, data on CRP as a triage test among all people with presumptive TB are limited and findings inconsistent. A meta-analysis of prospective studies evaluating CRP as a triage test among outpatients with presumptive TB demonstrated variability in accuracy by geography and HIV status.^8^ Similar results were found in a multi-country study analyzing CRP from banked plasma, but a more recent study from South Africa reported similar accuracy by HIV status.^9,10^

To better characterize the utility of CRP as a TB triage test, we performed a prospective, multi-country study among outpatient adults with presumptive TB in five high TB burden settings in Uganda, South Africa, the Philippines, Vietnam, and India. Our objectives were to 1) determine whether the sensitivity and specificity of CRP, measured at the point-of-care from capillary blood, meets or exceeds minimum accuracy targets for a TB triage test; 2) determine whether CRP performance differs by country, HIV status, diabetes status and/or sex and, 3) identify factors that most impact CRP performance.

## Methods

### Study Participants

We screened adults (age ≥18 years) presenting to outpatient clinics in Kampala, Uganda; Cape Town, South Africa; Desmariñas City, Philippines; Hanoi, Vietnam; and Vellore, India for new or worsening cough ≥2 weeks. We excluded people who received treatment for TB infection or disease in the prior 12 months, took other medications with anti-mycobacterial activity (*e.g.,* fluoroquinolones) in the past 2 weeks, lived more than 20 km from the study site or expressed unwillingness to return for follow-up.

### Procedures

Trained study personnel collected demographic and clinical data using a standardized questionnaire and assessed participants for presence and duration of TB symptoms. CRP concentrations were measured at enrollment from whole blood obtained from fingerprick using a US Food and Drug Administration-approved standard sensitivity point-of-care assay (ichroma CRP rapid test, Boditech, South Korea) and a dedicated fluorescence-based immunoassay analyzer (ichroma^TM^ II, Boditech, South Korea) that provided quantitative CRP results (range: 2.5-300 mg/L) in three minutes. Participants provided two to three spot sputum specimens (≥1 mL each; induced with nebulized 3%-10% NaCl solution, if necessary) upon study entry for reference standard TB testing by trained laboratory technicians blinded to clinical and demographic data (including CRP levels), in quality-assured mycobacteriology laboratories. TB testing included Xpert MTB/RIF Ultra (Ultra; Cepheid, CA, USA), inclusive of repeat testing if the initial result was trace or invalid, and liquid mycobacterial culture x2 (for participants without positive Ultra results), as described previously.^11^

### Definitions

We defined *a priori* a point-of-care CRP concentration of ≥5 mg/L as a positive result in accordance with WHO guidelines for TB screening among PWH.^2^ We considered participants to be “TB-positive” by our microbiologic reference standard (MRS) if they had ≥1 positive sputum Xpert result (defined as semi-quantitative result of “very low” or higher), two “trace” sputum Xpert results, or *M. tuberculosis* isolated from ≥1 sputum culture. We required participants to have two negative culture results (and not meet the criteria above) to be “TB-negative”. Participants with insufficient data to classify TB status were excluded from analysis. To assess performance of CRP triage testing in routine settings where culture is typically not available, we also evaluated CRP accuracy in reference to sputum Xpert results (SXRS) alone.

### Statistical Analysis

A statistical analysis plan including reference standard definitions was finalized prior to analysis. We planned for a minimum sample size of 1500 participants (N=300 per country/year) such that the width of the 95% confidence interval (CI) would be less than ±9.4% if test sensitivity was ≥80% and less than ±5.6% if observed test specificity was ≥60%, assuming a TB prevalence of 20%. To enable sub-group analyses, we continued enrollment beyond our planned sample size of 1500 participants.

We summarized categorical and continuous variables for enrolled participants. We calculated the point estimates and 95% confidence intervals (CIs) for the sensitivity and specificity of CRP (using a 5 mg/L cut-point) using both reference standards. We used Chi-squared tests to compare differences in sensitivity and specificity between countries and pre-defined sub-groups including PWH, people living with diabetes (PWD), and by sex. We performed receiver operating characteristic (ROC) curve analysis and calculated area under the curve (AUC) with 95% CIs overall, by country and for each sub-group. We used De Long’s method to compare AUC between countries and between sub-groups. To identify factors impacting AUC, we built a parametric multivariate ROC regression model using maximum likelihood estimation. We included variables selected *a priori* (age, sex, country, HIV positive with CD4 count ≤200, diabetes, measures of TB disease severity) and additional variables with p<0.2 in the unadjusted models. For each variable, we evaluated the magnitude and direction (positive or negative) of the beta-coefficients in order to identify the variables having the greatest influence on CRP performance. We also used tobit regression to assess whether any demographic or clinical characteristics were associated with elevated CRP. Analyses were conducted using Stata Version 17 (StataCorp, College Station, Texas, USA).

### Ethics

The study was approved by institutional review boards at the University of California San Francisco (USA), University of Heidelberg (Germany), Makerere University (Uganda), Stellenbosch University (South Africa), Christian Medical College (India), National Lung Hospital (Vietnam) and De La Salle Medical and Health Sciences Institute (Philippines). This manuscript adheres to guidelines for reporting of diagnostic accuracy studies.^12^

## Results

### Study population/Patient demographic and clinical characteristics

From April 2021 to October 2023, we screened 4264 outpatient adults with cough ≥2 weeks and enrolled 3232 (76%) meeting study eligibility criteria (**Figure 1**). Of these, we excluded 105 (3%) participants for whom CRP testing was not performed and 223 (7%) participants with missing or incomplete TB test results from the analysis. Baseline characteristics of the remaining 2904 varied across countries (**Table 1**). The proportions of individuals reporting a prior history of TB ranged from 13% in Uganda to 31% in South Africa, with HIV ranged from 1% in Vietnam to 38% in South Africa, with diabetes ranged from 5% in South Africa to 22% in India (**Supplementary Table S1** describes baseline diabetes characteristics of PWD). The proportion of individuals with elevated CRP concentrations (≥5 mg/L) was lowest (26%) in the Philippines and highest (67%) in South Africa, and median CRP concentration was lowest (2.5 mg/L) in the Philippines and highest (10.7 mg/L) in Uganda (**Table 1**).

**Figure 1.**
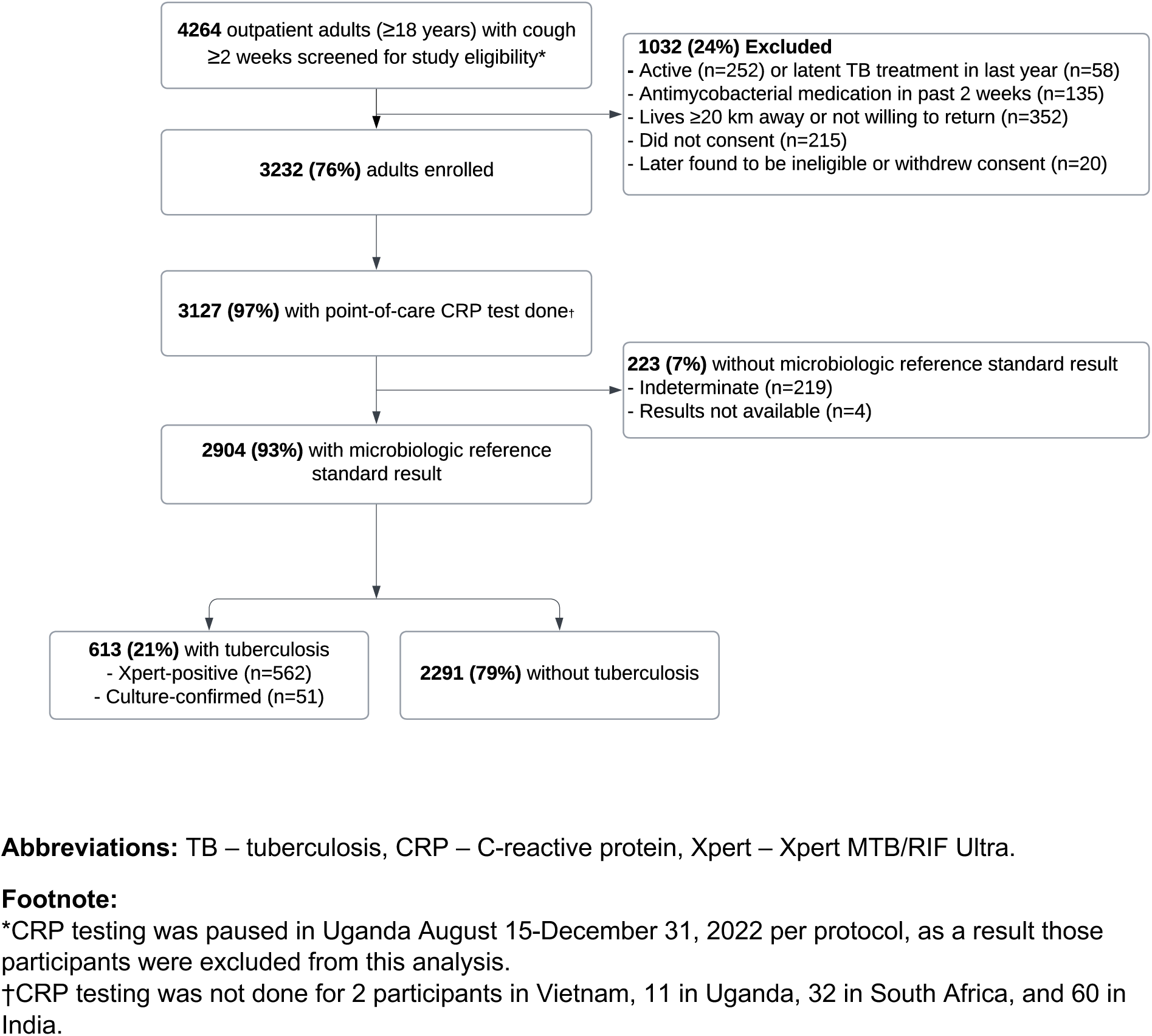
Participant flow chart.

**Table 1:**
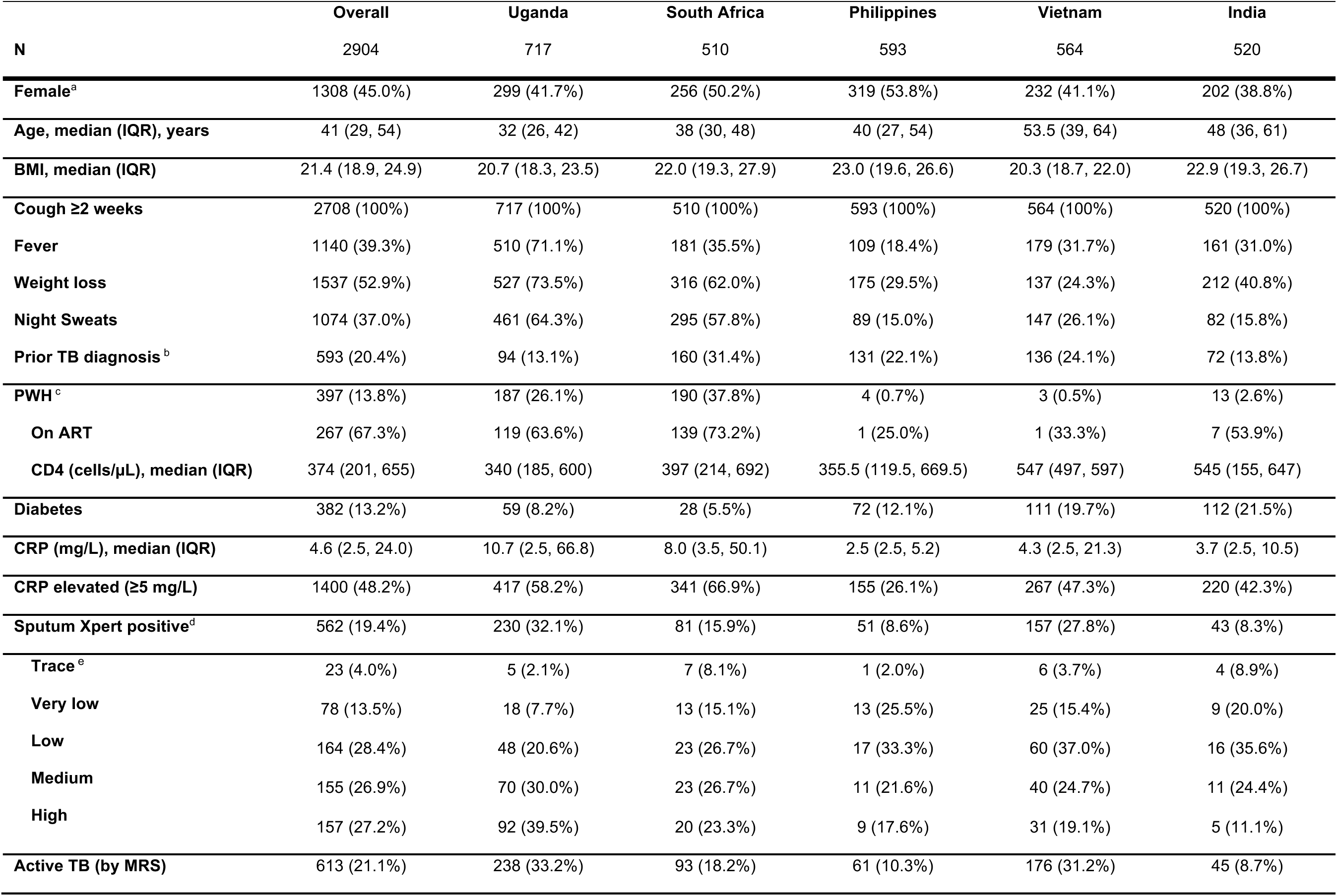
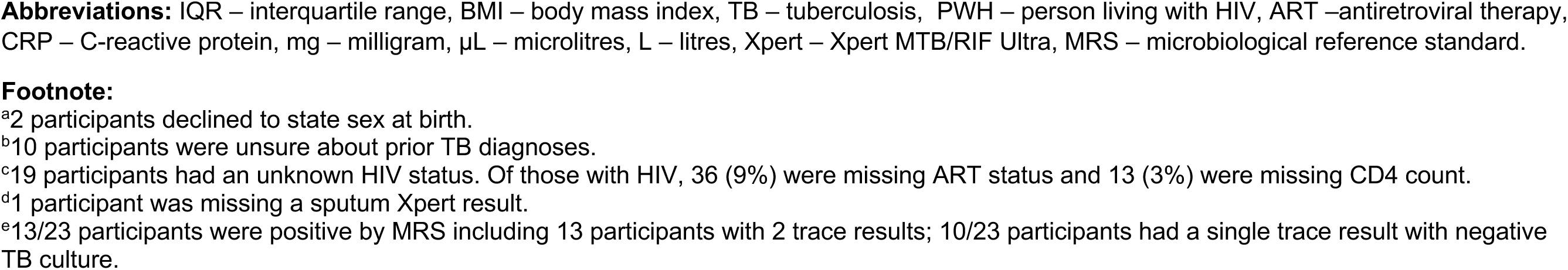
Demographical and clinical characteristics overall and by country.

|  | **Overall** | **Uganda** | **South Africa** | **Philippines** | **Vietnam** | **India** |
| --- | --- | --- | --- | --- | --- | --- |
| **N** | 2904 | 717 | 510 | 593 | 564 | 520 |
| **Female<sup>a</sup>** | 1308 (45.0%) | 299 (41.7%) | 256 (50.2%) | 319 (53.8%) | 232 (41.1%) | 202 (38.8%) |
| **Age, median (IQR), years** | 41 (29, 54) | 32 (26, 42) | 38 (30, 48) | 40 (27, 54) | 53.5 (39, 64) | 48 (36, 61) |
| **BMI, median (IQR)** | 21.4 (18.9, 24.9) | 20.7 (18.3, 23.5) | 22.0 (19.3, 27.9) | 23.0 (19.6, 26.6) | 20.3 (18.7, 22.0) | 22.9 (19.3, 26.7) |
| **Cough ≥2 weeks** | 2708 (100%) | 717 (100%) | 510 (100%) | 593 (100%) | 564 (100%) | 520 (100%) |
| **Fever** | 1140 (39.3%) | 510 (71.1%) | 181 (35.5%) | 109 (18.4%) | 179 (31.7%) | 161 (31.0%) |
| **Weight loss** | 1537 (52.9%) | 527 (73.5%) | 316 (62.0%) | 175 (29.5%) | 137 (24.3%) | 212 (40.8%) |
| **Night Sweats** | 1074 (37.0%) | 461 (64.3%) | 295 (57.8%) | 89 (15.0%) | 147 (26.1%) | 82 (15.8%) |
| **Prior TB diagnosis<sup>b</sup>** | 593 (20.4%) | 94 (13.1%) | 160 (31.4%) | 131 (22.1%) | 136 (24.1%) | 72 (13.8%) |
| **PWH<sup>c</sup>** | 397 (13.8%) | 187 (26.1%) | 190 (37.8%) | 4 (0.7%) | 3 (0.5%) | 13 (2.6%) |
| **On ART** | 267 (67.3%) | 119 (63.6%) | 139 (73.2%) | 1 (25.0%) | 1 (33.3%) | 7 (53.9%) |
| **CD4 (cells/μL), median (IQR)** | 374 (201, 655) | 340 (185, 600) | 397 (214, 692) | 355.5 (119.5, 669.5) | 547 (497, 597) | 545 (155, 647) |
| **Diabetes** | 382 (13.2%) | 59 (8.2%) | 28 (5.5%) | 72 (12.1%) | 111 (19.7%) | 112 (21.5%) |
| **CRP (mg/L), median (IQR)** | 4.6 (2.5, 24.0) | 10.7 (2.5, 66.8) | 8.0 (3.5, 50.1) | 2.5 (2.5, 5.2) | 4.3 (2.5, 21.3) | 3.7 (2.5, 10.5) |
| **CRP elevated (≥5 mg/L)** | 1400 (48.2%) | 417 (58.2%) | 341 (66.9%) | 155 (26.1%) | 267 (47.3%) | 220 (42.3%) |
| **Sputum Xpert positive<sup>d</sup>** | 562 (19.4%) | 230 (32.1%) | 81 (15.9%) | 51 (8.6%) | 157 (27.8%) | 43 (8.3%) |
| **Trace<sup>e</sup>** | 23 (4.0%) | 5 (2.1%) | 7 (8.1%) | 1 (2.0%) | 6 (3.7%) | 4 (8.9%) |
| **Very low** | 78 (13.5%) | 18 (7.7%) | 13 (15.1%) | 13 (25.5%) | 25 (15.4%) | 9 (20.0%) |
| **Low** | 164 (28.4%) | 48 (20.6%) | 23 (26.7%) | 17 (33.3%) | 60 (37.0%) | 16 (35.6%) |
| **Medium** | 155 (26.9%) | 70 (30.0%) | 23 (26.7%) | 11 (21.6%) | 40 (24.7%) | 11 (24.4%) |
| **High** | 157 (27.2%) | 92 (39.5%) | 20 (23.3%) | 9 (17.6%) | 31 (19.1%) | 5 (11.1%) |
| **Active TB (by MRS)** | 613 (21.1%) | 238 (33.2%) | 93 (18.2%) | 61 (10.3%) | 176 (31.2%) | 45 (8.7%) |

**Abbreviations:** IQR – interquartile range, BMI – body mass index, TB – tuberculosis, PWH – person living with HIV, ART –antiretroviral therapy, CRP – C-reactive protein, mg – milligram, µL – microlitres, L – litres, Xpert – Xpert MTB/RIF Ultra, MRS – microbiological reference standard.
<sup>a</sup>2 participants declined to state sex at birth.
<sup>b</sup>10 participants were unsure about prior TB diagnoses.
<sup>c</sup>19 participants had an unknown HIV status. Of those with HIV, 36 (9%) were missing ART status and 13 (3%) were missing CD4 count.
<sup>d</sup>1 participant was missing a sputum Xpert result.
<sup>e</sup>13/23 participants were positive by MRS including 13 participants with 2 trace results; 10/23 participants had a single trace result with negative TB culture.

Overall, 613 (21%) patients had confirmed TB by the MRS including 562 (92%) with positive sputum Xpert results and 51 (8%) with positive sputum culture results only. TB prevalence varied across countries; prevalence was lowest in India (9%) and highest in Uganda (33%).

### Diagnostic accuracy of CRP overall and by country

When compared to MRS, the AUC for CRP was 0.81 (95% CI: 0.79-0.83) (**Figure 2**). We observed higher AUC for CRP (0.82-0.83) for African countries and lower AUC (0.76-0.80) for Asian countries.

**Figure 2.**
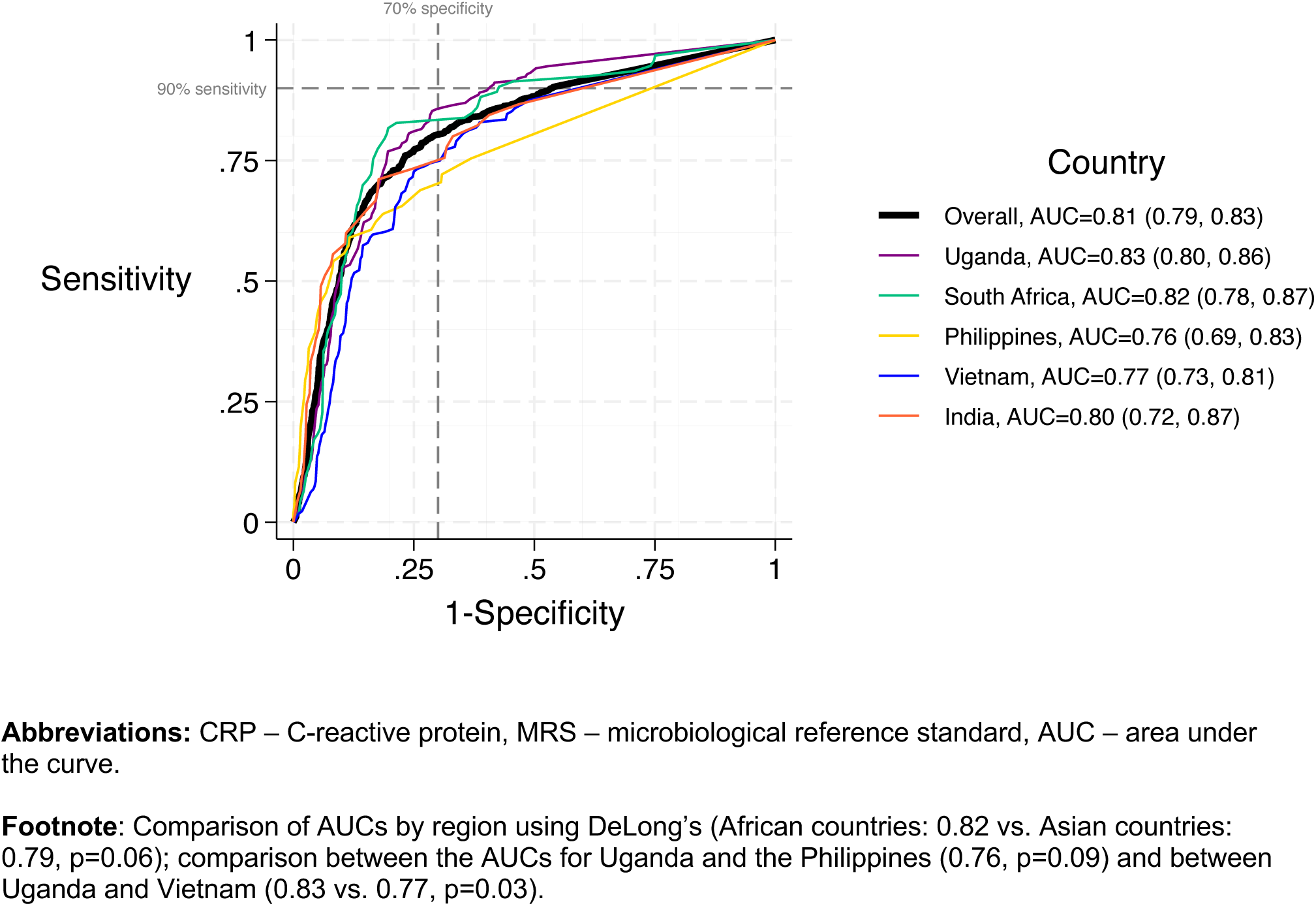
Receiver operating characteristic analysis for CRP in reference to MRS, overall study population and by country.

Using the WHO-recommended cut-point (≥5 mg/L), sensitivity of CRP was 84% (95% CI: 81-87%) and specificity was 61% (95% CI: 59-63%) (**Table 2**). Sensitivity was highest in African countries (91-93%) and lowest in Asian countries (64-82%) (**Table S2**, p<0.01). Specificity was highest in Asian countries (62-78%), exceeding 70% in the Philippines only, and lowest in African countries (39-58%) (p<0.01). Sensitivity and specificity estimates were similar overall and for each country when using sputum Xpert results alone as the reference standard (**Table S3**).

**Table 2.**
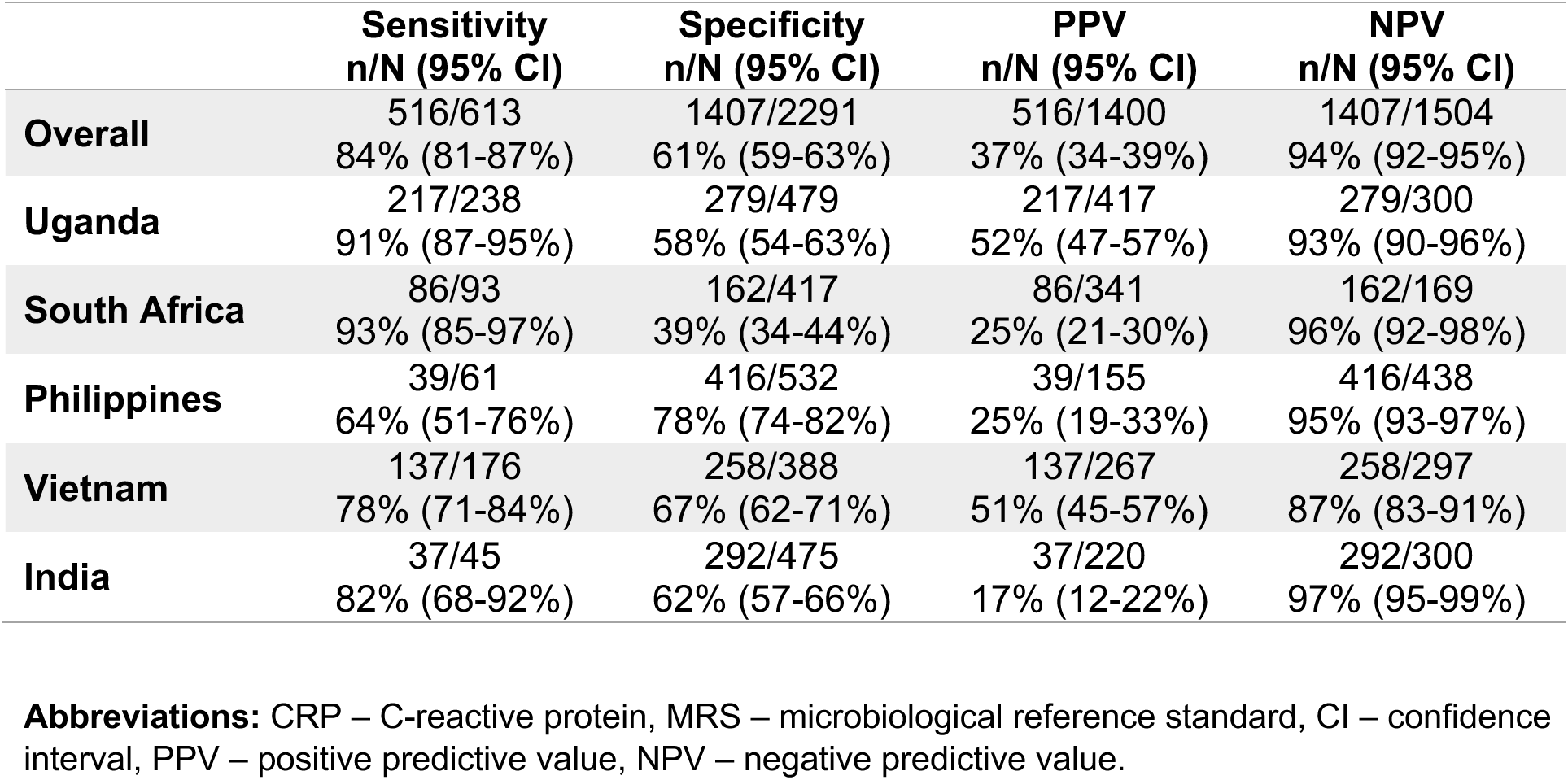
Diagnostic accuracy of CRP at cut-off point of 5 mg/L in reference to MRS, overall study population and by country.

|  | **Sensitivity**<br>**n/N (95% CI)** | **Specificity**<br>**n/N (95% CI)** | **PPV**<br>**n/N (95% CI)** | **NPV**<br>**n/N (95% CI)** |
| --- | --- | --- | --- | --- |
| **Overall** | 516/613<br>84% (81-87%) | 1407/2291<br>61% (59-63%) | 516/1400<br>37% (34-39%) | 1407/1504<br>94% (92-95%) |
| **Uganda** | 217/238<br>91% (87-95%) | 279/479<br>58% (54-63%) | 217/417<br>52% (47-57%) | 279/300<br>93% (90-96%) |
| **South Africa** | 86/93<br>93% (85-97%) | 162/417<br>39% (34-44%) | 86/341<br>25% (21-30%) | 162/169<br>96% (92-98%) |
| **Philippines** | 39/61<br>64% (51-76%) | 416/532<br>78% (74-82%) | 39/155<br>25% (19-33%) | 416/438<br>95% (93-97%) |
| **Vietnam** | 137/176<br>78% (71-84%) | 258/388<br>67% (62-71%) | 137/267<br>51% (45-57%) | 258/297<br>87% (83-91%) |
| **India** | 37/45<br>82% (68-92%) | 292/475<br>62% (57-66%) | 37/220<br>17% (12-22%) | 292/300<br>97% (95-99%) |

**Abbreviations:** CRP – C-reactive protein, MRS – microbiological reference standard, CI – confidence interval, PPV – positive predictive value, NPV – negative predictive value.

### Diagnostic accuracy of CRP in sub-populations

**Figure 3** shows the ROC for CRP in reference to MRS overall and for key sub-groups (PWH, PWD) and by sex. There was no difference in CRP accuracy by HIV status, diabetes status, or sex (AUC comparison p-value ≥0.10 for all 3 sub-groups**, Table S4**). **Supplementary tables S5, S6 and S7** describe the sensitivity and specificity of CRP in reference to MRS and SXRS, by HIV status, diabetes status, and sex, respectively. At a 5 mg/L cut-point, CRP did not achieve the minimum 90% target for sensitivity or 70% target for specificity in any sub-population in reference to MRS but sensitivity reached 90% among PWH in reference to sputum Xpert results alone. Among PWD, sensitivity of CRP exceeded 90% among certain diabetes sub-groups, including those with newly diagnosed diabetes and those with HbA1c <7% (**Table S8**).

**Figure 3.**
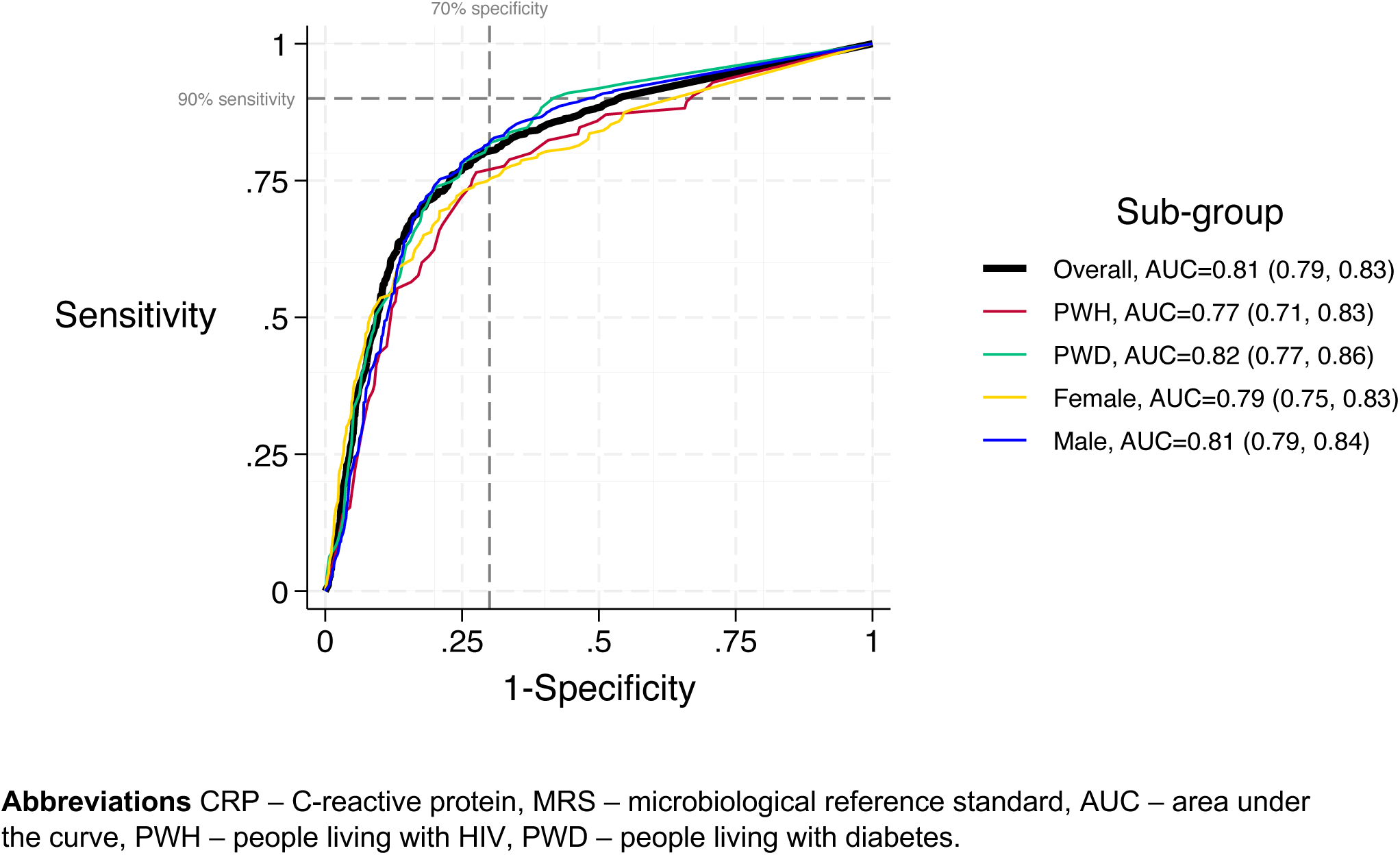
Receiver operating characteristic analysis for CRP in reference to MRS, overall study population and by key sub-groups.

CRP sensitivity was higher in men than women (87% vs. 79%, difference +8% [95% CI: 1 to 15%], p=0.02) and specificity was higher in people without HIV than PWH (64% vs. 45%, difference +19% [95% CI: 13 to 25%], p<0.01). There was no evidence of a difference in sensitivity and specificity by any other sub-group.

### ROC regression

In univariable models, country, multiple markers of disease or illness severity (HIV-positive with CD4 count ≤200, heart rate [HR], weight loss, and body mass index [BMI]) and a marker of TB disease severity (Xpert semi-quantitative grade) were associated with the diagnostic accuracy of CRP. In the multivariable model, only country, HIV-positive with CD4 count ≤200, HR, BMI and Xpert semi-quantitative grade were significantly associated with CRP accuracy (p<0.05, **Table S9**), of which, beta-coefficients (absolute value) were largest for the following variables: PWH with CD4 count **≤**200, country and Xpert semi-quantitative grade. The direction of the beta-coefficient indicated the directional impact of the variable on CRP accuracy. Asian countries were associated with reduced CRP accuracy, as evidenced by negative beta-coefficients (Philippines B=-0.38, 95% CI: -0.71 to -0.04 and Vietnam B=-0.32, 95% CI: -0.60 to -0.05). High HR (B=0.01, 95% CI: +0.002 to +0.013), low BMI (B=0.04, 95% CI: +0.02 to +0.07), and PWH with CD4 count **≤**200 (B=0.43, 95% CI: +0.02 to +0.84) were associated with increased CRP accuracy. Among those with active TB, higher mycobacterial burden, which was more common among African countries, increased CRP accuracy (medium/high Xpert semi-quantitative grade B=0.44, 95% CI: +0.27 to +0.62). Other variables such as diabetes was associated with elevated CRP levels in tobit regression (**Table S10**) but was not associated with change in CRP accuracy in the ROC regression (**Table S9**).

## Discussion

CRP was recently endorsed by the WHO as a new TB screening tool for PWH, based mostly on data from sub-Saharan Africa.^2,7^ However, the usefulness of CRP for TB triage testing among symptomatic individuals with and without HIV and from other settings is unknown. In this multi-country evaluation among outpatient adults with cough ≥2 weeks, we found that CRP did not meet the minimum sensitivity (90%) or specificity targets (70%) recommended by the WHO for an effective TB triage test overall (sensitivity 84% and specificity 63%, in reference to MRS). Although CRP performed better in some countries and sub-groups, there was no country or sub-group where CRP met the minimum targets for both sensitivity and specificity. These results suggest that CRP is unlikely to be broadly useful for TB triage testing.

Our study is the first multi-country prospective evaluation of CRP-based TB triage testing and the first study to evaluate the diagnostic accuracy of CRP using the WHO-recommended 5 mg/L cut-point, *a priori*. To-date, prior studies evaluating CRP TB triage testing have been limited to case-control studies and one prospective study.^9,10^ We found that while CRP did not meet the minimum WHO targets for sensitivity or specificity overall, CRP demonstrated substantial geographic variability such that sensitivity exceeded 91% in reference to MRS in Africa but was much lower (64-82%) in Asia, where specificity was highest. These geographic differences in CRP sensitivity, which have also been reported by prior studies, correlated with observed geographic differences in mycobacterial load.^8,9,13^ Consistent with the prior multi-national study using stored specimens^9^, CRP sensitivity in our study was lower in Asia (where 36-44% of Xpert-positive participants had a semi-quantitative result of “medium” or “high”) than in Africa (where 50-70% of Xpert-positive participants had a semi-quantitative result of “medium” or “high”). These results suggest that factors affecting mycobacterial load in a population are likely to have substantial impact on CRP accuracy.

The diagnostic accuracy of any test is sensitive to differences in the characteristics of the population in which the test is evaluated, particularly differences in the spectrum of disease severity. For TB, the spectrum of clinical disease severity is associated with mycobacterial load^14–19^, and multiple measurements of mycobacterial load (*e.g.,* smear status, Xpert semi-quantitative grade, time-to-culture positivity) have been correlated with CRP levels.^9,20–24^ Other patient factors (*e.g.,* biologic characteristics, behaviors) and health system factors can influence the mycobacterial load in a population. Biologic characteristics such as HIV and diabetes are known to affect mycobacterial load and therefore TB test accuracy by influencing disease susceptibility and severity.^25,26^ Mycobacterial load is also susceptible to factors leading to delays in TB diagnosis, such as patient health-seeking behavior and accessibility of TB diagnostic services. To better understand the factors affecting the diagnostic accuracy of CRP, we evaluated CRP accuracy by HIV status, diabetes status and sex. Additionally, we performed a ROC regression to identify additional factors, including measures of mycobacterial load, that may contribute to differences in CRP performance.

When we evaluated the diagnostic accuracy of CRP triage testing by HIV status, we found no difference in sensitivity in reference to both MRS and SXRS by HIV status but found that specificity was substantially lower among PWH relative to people without HIV (-19% difference in reference to MRS). Our results differ from an earlier study that enrolled more immunosuppressed PWH (lower median CD4 counts, smaller proportion of PWH on ART) and found HIV infection to be associated with increased CRP sensitivity.^9^ However, our results are similar to a more recent study with similar demographics for PWH as our study (high median CD4 count and high proportion of PWH on ART) which also found no difference in CRP sensitivity by HIV status.^10^ Combined, these studies suggest that HIV infection may have less of an impact on CRP performance when there are less differences in TB risk between people with and without HIV (*i.e.,* due to less immunosuppression among PWH) and may explain why HIV infection with CD4 count >200 was not associated with CRP accuracy in our ROC regression. As median pre-ART CD4 count and ART coverage increases, fewer settings are likely to report differences in CRP sensitivity by HIV status but differences in specificity may persist due to the increased susceptibility to other non-TB diseases associated with HIV infection (*e.g.,* bacterial pneumonia), regardless of CD4 count.

Alternatively, for people with known HIV infection, CRP could be used to screen unselected PWH or PWH who screen-positive by screening strategies more sensitive than cough ≥2 weeks (*i.e.,* WHO 4-part symptom screen, CXR), and in these scenarios, CRP can be expected to have higher specificity than reported here.^7,20^

Like HIV, diabetes is also associated with increased susceptibility to infections including pulmonary TB and more severe TB disease.^27,28^ However, in our study we found no differences in CRP sensitivity or specificity by diabetes status, perhaps due to the modest effect of diabetes on infectious disease risk, and no difference in CRP accuracy among different diabetic sub-groups. Larger studies are needed to better understand factors that may impact CRP performance among PWD. When we compared CRP accuracy by sex, we found that sensitivity was significantly higher among men (+8% difference in reference to MRS) but still failed to reach the minimum 90% sensitivity target when either MRS or SXRS was used as the reference standard. The higher sensitivity of CRP among men could at least partly be explained by sex differences in mycobacterial load, which many studies have largely attributed to delayed healthcare-seeking behavior.^29,30^ For men and people living in areas where time-to-TB diagnosis is delayed (populations expected to have higher mycobacterial load at diagnosis), our results suggest that CRP triage testing can be expected to detect the majority of those individuals with active TB who are at highest risk for poor outcomes from later TB diagnosis. However, a greater number of confirmatory tests may need to be performed per TB case detected, as the prevalence of non-TB disease causing elevations in CRP is also likely increased. In such settings or populations, additional triage testing (*e.g.,* CXR) may help improve overall specificity.

Conversely, our results also suggests that CRP triage testing is likely to miss a greater proportion of individuals with active TB in areas where the spectrum of clinical disease severity is milder and mycobacterial load is lower at diagnosis. Sensitivity of CRP was lowest in the Philippines, where TB appeared to be diagnosed at an earlier stage than other countries; participants in the Philippines with active TB reported the fewest number of symptoms, had the lowest median CRP levels and lowest mycobacterial load. Although study procedures were the same for all sites, some settings may be diagnosing TB earlier than other settings by, for example, routinely screening individuals for active TB or ensuring most of the population has access to high quality TB diagnostic services. In such settings, more sensitive TB diagnostics will be needed to increase TB case detection.

Our study has several strengths. First, ours is the largest study to-date to report the diagnostic accuracy of CRP as a triage test for pulmonary TB and our consecutive cohort of outpatients enrolled from five TB endemic countries with diverse TB, HIV and diabetes epidemiology ensures the generalizability of our findings. Second, diagnostic accuracy of CRP was evaluated in reference to both a robust microbiologic and a pragmatic reference standard that is routinely used in most high burden settings. Third, CRP concentrations were measured using a commercially available, simple, and low-cost point-of-care assay. As such, CRP is available for immediate scale-up for clinics wishing to implement CRP-based TB triage testing and further strengthen its evidence base.

Our study also has important limitations. First, we did not assess all variables that may contribute to differences in mycobacterial load and thus, CRP performance. Although prior studies have confirmed the association of certain patient and health system factors on measures of mycobacterial load ^31–33^ and, separately, studies have correlated mycobacterial load with CRP levels, ^9,20–24^ future studies should consider including variables that affect mycobacterial load in their data collection to more directly evaluate the extent to which these factors affect test performance. Second, we screened all prospective participants for cough ≥2 weeks, including PWH. Future studies should evaluate the performance and yield of CRP-based TB triage testing in combination with more sensitive screening tools such as the WHO 4-part symptom screen and CXR. Third, we identified individuals with active TB by evaluating sputum only and did not perform additional testing to evaluate for extrapulmonary TB or non-TB disease; studies assessing the causes and significance of elevated CRP levels among individuals with non-TB disease are particularly needed. Additionally, future studies should consider evaluating the impact of *Mtb* strain-type on TB test performance, as one prior study found an association between *Mtb* strain-type and host CRP response to TB.^13^

In conclusion, CRP in our study was not the one-size-fits-all TB triage test clinicians, researchers, and policymakers alike, were hoping for. We found that CRP performance varied greatly by setting and in some sub-groups, most likely reflecting population differences in mycobacterial load and the spectrum of clinical disease severity. More sensitive and specific tests with similar operational characteristics to CRP are needed for scale-up of TB triage testing in TB endemic settings.

## Data Availability

All data produced in the present study are available upon reasonable request to the authors

## Supplement

This article has an online data supplement (10 Supplementary Tables).

## Conflict of interest

William Worodria, Charles Yu, Nhung Viet Nguyen, Devasahayam Jesudas Christopher, Grant Theron, Patrick P.J. Philips, Payam Nahid, Claudia M. Denkinger, Adithya Cattamanchi, and Christina Yoon declare support from the underlying R2D2 TB Network to their institutions from the National Institute of Allergy and Infectious Diseases (NIAID) of the US National Institutes of Health (NIH). CMD also declares research grants from the German Ministry of Education and Research, German Alliance for Global Health Research, US Agency for International Development, FIND, German Center for Infection Research, and WHO. AC declares research funding to his institution from NIH, Bill and Melinda Gates Foundation, and Global Health Labs. CY declares research grants from NIH/NIAID and NIH/National Heart Lung Blood Institute, with CRP test strips and analyzers donated by Boditech Med Inc, South Korea. CY serves as a a Scientific Advisory Board member for an EDCTP-funded cluster randomized trial. All other authors declare no competing interests.

CRP test strips and analyzer were donated by Boditech Med Inc, South Korea.

## References

1. Global Tuberculosis Report 2023 [Internet]. [cited 2024 Feb 2]. Available from: https://www.who.int/teams/global-tuberculosis-programme/tb-reports/global-tuberculosis-report-2023

2. WHO consolidated guidelines on tuberculosis: module 3: diagnosis: rapid diagnostics for tuberculosis detection, 2021 update [Internet]. [cited 2022 Nov 21]. Available from: https://www.who.int/publications-detail-redirect/9789240029415

3. Hoog AH van’t, Cobelens F, Vassall A, Kampen S van, Dorman SE, Alland D, et al. Optimal Triage Test Characteristics to Improve the Cost-Effectiveness of the Xpert MTB/RIF Assay for TB Diagnosis: A Decision Analysis. PLOS ONE. 2013 Dec 18;8(12):e82786.

4. Jaeger S, Karargyris A, Candemir S, Folio L, Siegelman J, Callaghan F, et al. Automatic tuberculosis screening using chest radiographs. IEEE Trans Med Imaging. 2014 Feb;33(2):233–45.

5. Murphy K, Habib SS, Zaidi SMA, Khowaja S, Khan A, Melendez J, et al. Computer aided detection of tuberculosis on chest radiographs: An evaluation of the CAD4TB v6 system. Sci Rep. 2020 Mar 26;10(1):5492.

6. Soares TR, Oliveira RD de, Liu YE, Santos A da S, Santos PCP dos, Monte LRS, et al. Evaluation of chest X-ray with automated interpretation algorithms for mass tuberculosis screening in prisons: a cross-sectional study. Lancet Reg Health Am. 2022 Nov 4;17:100388.

7. Dhana A, Hamada Y, Kengne AP, Kerkhoff AD, Rangaka MX, Kredo T, et al. Tuberculosis screening among ambulatory people living with HIV: a systematic review and individual participant data meta-analysis. The Lancet Infectious Diseases. 2022 Apr 1;22(4):507–18.

8. Santos VS, Goletti D, Kontogianni K, Adams ER, Molina-Moya B, Dominguez J, et al. Acute phase proteins and IP-10 as triage tests for the diagnosis of tuberculosis: systematic review and meta-analysis. Clinical Microbiology and Infection. 2019 Feb 1;25(2):169–77.

9. Samuels THA, Wyss R, Ongarello S, Moore DAJ, Schumacher SG, Denkinger CM. Evaluation of the diagnostic performance of laboratory-based c-reactive protein as a triage test for active pulmonary tuberculosis. PLoS One. 2021;16(7):e0254002.

10. Calderwood CJ, Reeve BW, Mann T, Palmer Z, Nyawo G, Mishra H, et al. Clinical utility of C-reactive protein-based triage for presumptive pulmonary tuberculosis in South African adults. J Infect. 2023 Jan;86(1):24–32.

11. Gupta-Wright A, Ha H, Abdulgadar S, Crowder R, Emmanuel J, Mukwatamundu J, et al. Evaluation of the Xpert MTB Host Response assay for the triage of patients with presumed pulmonary tuberculosis: a prospective diagnostic accuracy study in Viet Nam, India, the Philippines, Uganda, and South Africa. The Lancet Global Health. 2024 Feb 1;12(2):e226–34.

12. Bossuyt PM, Reitsma JB, Bruns DE, Gatsonis CA, Glasziou PP, Irwig L, et al. STARD 2015: an updated list of essential items for reporting diagnostic accuracy studies. BMJ. 2015 Oct 28;351:h5527.

13. Brown J, Clark K, Smith C, Hopwood J, Lynard O, Toolan M, et al. Variation in C - reactive protein response according to host and mycobacterial characteristics in active tuberculosis. BMC Infectious Diseases. 2016 Jun 10;16(1):265.

14. Beynon F, Theron G, Respeito D, Mambuque E, Saavedra B, Bulo H, et al. Correlation of Xpert MTB/RIF with measures to assess Mycobacterium tuberculosis bacillary burden in high HIV burden areas of Southern Africa. Sci Rep. 2018 Mar 26;8(1):5201.

15. Becerra MC, Pachao-Torreblanca IF, Bayona J, Celi R, Shin SS, Kim JY, et al. Expanding tuberculosis case detection by screening household contacts. Public Health Rep. 2005;120(3):271–7.

16. Perrin FMR, Woodward N, Phillips PPJ, McHugh TD, Nunn AJ, Lipman MCI, et al. Radiological cavitation, sputum mycobacterial load and treatment response in pulmonary tuberculosis. The International Journal of Tuberculosis and Lung Disease. 2010 Dec 1;14(12):1596–602.

17. Fortún J, Martín-Dávila P, Molina A, Navas E, Hermida JM, Cobo J, et al. Sputum conversion among patients with pulmonary tuberculosis: are there implications for removal of respiratory isolation? Journal of Antimicrobial Chemotherapy. 2007 Apr 1;59(4):794–8.

18. Bark CM, Thiel BA, Johnson JL. Pretreatment Time to Detection of Mycobacterium tuberculosis in Liquid Culture Is Associated with Relapse after Therapy. J Clin Microbiol. 2012 Feb;50(2):538.

19. Palaci M, Dietze R, Hadad DJ, Ribeiro FKC, Peres RL, Vinhas SA, et al. Cavitary Disease and Quantitative Sputum Bacillary Load in Cases of Pulmonary Tuberculosis. J Clin Microbiol. 2007 Dec;45(12):4064–6.

20. Yoon C, Semitala FC, Atuhumuza E, Katende J, Mwebe S, Asege L, et al. Point-of-care C-reactive protein-based tuberculosis screening for people living with HIV: a diagnostic accuracy study. Lancet Infect Dis. 2017 Dec;17(12):1285–92.

21. Lawn SD, Kerkhoff AD, Vogt M, Wood R. Diagnostic and prognostic value of serum C-reactive protein for screening for HIV-associated tuberculosis. Int J Tuberc Lung Dis. 2013 May;17(5):10.5588/ijtld.12.0811.

22. Ciccacci F, Floridia M, Bernardini R, Sidumo Z, Mugunhe RJ, Andreotti M, et al. Plasma levels of CRP, neopterin and IP-10 in HIV-infected individuals with and without pulmonary tuberculosis. J Clin Tuberc Other Mycobact Dis. 2019 Jun 5;16:100107.

23. Shameem M, Fatima N, Ahmad A, Malik A, Husain Q. Correlation of Serum C-Reactive Protein with Disease Severity in Tuberculosis Patients. Open Journal of Respiratory Diseases. 2012 Nov 6;2(4):95–100.

24. Rao S, Bernhardt V. Serum C-Reactive Protein in Pulmonary Tuberculosis: Correlation With Bacteriological Load and Extent of Disease. Infectious Diseases in Clinical Practice. 2009 Sep;17(5):314.

25. Lawn SD, Churchyard G. Epidemiology of HIV-associated tuberculosis Running Head: Epidemiology of TB /HIV. Curr Opin HIV AIDS. 2009 Jul;4(4):325–33.

26. Harries AD, Satyanarayana S, Kumar AMV, Nagaraja SB, Isaakidis P, Malhotra S, et al. Epidemiology and interaction of diabetes mellitus and tuberculosis and challenges for care: a review. Public Health Action. 2013 Nov 4;3(Suppl 1):S3–9.

27. Ronacher K, Joosten SA, van Crevel R, Dockrell HM, Walzl G, Ottenhoff THM. Acquired immunodeficiencies and tuberculosis: focus on HIV/AIDS and diabetes mellitus. Immunol Rev. 2015 Mar;264(1):121–37.

28. Jeon CY, Murray MB. Diabetes mellitus increases the risk of active tuberculosis: a systematic review of 13 observational studies. PLoS Med. 2008 Jul 15;5(7):e152.

29. Borgdorff MW, Nagelkerke NJ, Dye C, Nunn P. Gender and tuberculosis: a comparison of prevalence surveys with notification data to explore sex differences in case detection. Int J Tuberc Lung Dis. 2000 Feb;4(2):123–32.

30. Horton KC, MacPherson P, Houben RMGJ, White RG, Corbett EL. Sex Differences in Tuberculosis Burden and Notifications in Low- and Middle-Income Countries: A Systematic Review and Meta-analysis. PLoS Med. 2016 Sep;13(9):e1002119.

31. Virenfeldt J, Rudolf F, Camara C, Furtado A, Gomes V, Aaby P, et al. Treatment delay affects clinical severity of tuberculosis: a longitudinal cohort study. BMJ Open. 2014 Jun 10;4(6):e004818.

32. Paramasivam S, Thomas B, Chandran P, Thayyil J, George B, Sivakumar CP. Diagnostic delay and associated factors among patients with pulmonary tuberculosis in Kerala. J Family Med Prim Care. 2017;6(3):643–8.

33. Lawn SD, Afful B, Acheampong JW. Pulmonary tuberculosis: diagnostic delay in Ghanaian adults. Int J Tuberc Lung Dis. 1998 Aug;2(8):635–40.

