## Supplemental Tables for "C-reactive protein-based tuberculosis triage testing: a multi-country diagnostic accuracy study"

**Table S1. Descriptive characteristics of participants with diabetes.**

| | Overall N (%) | Uganda | South Africa | Philippines | Vietnam | India |
|---|---|---|---|---|---|---|
| **PWD** | 356/2708 (13%) | 58/709 (8%) | 23/424 (5%) | 72/582 (12%) | 107/544 (20%) | 96/449 (21%) |
| **Prior diabetes diagnosis** | 215 (60%) | 15 (26%) | 10 (44%) | 46 (64%) | 71 (66%) | 73 (76%) |
| **On insulin** | 24 (11%) | 2 (13%) | 3 (30%) | 1 (2%) | 11 (16%) | 7 (10%) |
| **HbA1c (%), median**[a] | 6.9 (6.4, 8.6) | 6.7 (6.5, 7.3) | 6.7 (6, 7.1) | 7.2 (6.1, 10.1) | 6.8 (6.2, 9.2) | 7.2 (6.5, 8.8) |
| **HbA1c ≥7.0%** | 170/352 (48%) | 23/58 (40%) | 9/22 (41%) | 40/72 (56%) | 46/107(43%) | 52/93 (56%) |

**Abbreviations:** PWD – people with diabetes, HbA1c – hemoglobin A1c.

**Footnote:** [a]Four participants were missing HbA1c result.

**Table S2. Diagnostic accuracy of CRP (5 mg/L cut-point) in reference to MRS, by region.**

| | Africa N=1227 | Asia N=1677 | Difference (95% CI) | P-value for difference |
|---|---|---|---|---|
| **Sensitivity n/N (95% CI)** | 303/331 92% (88-94%) | 213/282 76% (70-80%) | -16% (-22 to -10%) | <0.01 |
| **Specificity n/N (95% CI)** | 441/896 49% (46-53%) | 966/1395 69% (67-72%) | +20% (16 to 24%) | <0.01 |
| **PPV n/N (95% CI)** | 303/758 40% (37-44%) | 213/642 33% (30-37%) | -7% (-12 to 2%) | <0.01 |
| **NPV n/N (95% CI)** | 441/469 94% (92-96%) | 966/1035 93% (92-95%) | -1% (-3 to 2%) | 0.61 |

**Abbreviations:** CRP – C-reactive protein, MRS – microbiological reference standard, CI – confidence interval, PPV – positive predictive value, NPV – negative predictive value.

**Table S3. Diagnostic accuracy of CRP (5 mg/L cut-point) in reference to sputum Xpert MTB/RIF Ultra results, overall study population and by country.**

| | Sensitivity (95% CI) | Specificity (95% CI) | PPV (95% CI) | NPV (95% CI) |
|---|---|---|---|---|
| **Overall** | 487/562<br>87% (84-89%) | 1423/2325<br>61% (59-63%) | 487/1389<br>35% (33-38%) | 1423/1498<br>95% (94-96%) |
| **Uganda** | 211/230<br>92% (87-95%) | 280/484<br>58% (53-62%) | 211/415<br>51% (46-56%) | 280/299<br>94% (90-96%) |
| **South Africa** | 79/81<br>98% (91-100%) | 166/424<br>39% (35-44%) | 79/337<br>23% (19-28%) | 166/168<br>99% (96-100%) |
| **Philippines** | 36/51<br>71% (56-83%) | 423/542<br>78% (74-82%) | 36/155<br>23% (17-31%) | 423/438<br>97% (94-98%) |
| **Vietnam** | 126/157<br>80% (73-86%) | 263/402<br>65% (61-70%) | 126/265<br>48% (41-54%) | 263/294<br>90% (85-93%) |
| **India** | 35/43<br>81% (67-92%) | 291/473<br>62% (57-66%) | 35/217<br>16% (12-22%) | 291/299<br>97% (95-99%) |

**Abbreviations:** CRP – C-reactive protein, CI – confidence interval, PPV – positive predictive value, NPV – negative predictive value.

**Table S4. Comparison of sub-group AUCs using DeLong's method.**

| Sub-group | AUC (95% CI) | p-value |
|---|---|---|
| **PWH** | 0.77 (0.71, 0.83) | 0.10 |
| **People without HIV** | 0.82 (0.80, 0.84) | |
| **PWD** | 0.82 (0.77, 0.86) | 0.76 |
| **People without diabetes** | 0.81 (0.79, 0.83) | |
| **Male** | 0.81 (0.79, 0.84) | 0.27 |
| **Female** [a] | 0.79 (0.75, 0.83) | |

**Abbreviations:** AUC – area under the curve, CI – confidence interval, PWH – people living with HIV, PWD – people living with diabetes.

**Footnote:** [a]Two participants declined to state sex at birth.

**Table S5. Diagnostic accuracy of CRP (5 mg/L cut-point) in reference to MRS and SXRS, by HIV status.**

| | Overall | HIV status [a] | | | |
|---|---|---|---|---|---|
| **In reference to MRS** | | **PWH** | **HIV-negative** | **Difference (95% CI)** | **p-value** |
| **Sensitivity n/N (95% CI)** | 516/613<br>84% (81-87%) | 74/85<br>87% (78-93%) | 441/526<br>84% (80-87%) | -3%<br>(-11 to 5%) | 0.45 |
| **Specificity n/N (95% CI)** | 1407/2291<br>61% (59-63%) | 140/312<br>45% (39-51%) | 1260/1962<br>64% (62-66%) | +19%<br>(13 to 25%) | <0.01 |
| **In reference to SXRS** | | | | | |
| **Sensitivity n/N (95% CI)** | 487/562<br>87% (84-89%) | 64/71<br>90% (81-96%) | 423/490<br>86% (83-89%) | -4%<br>(-11 to 4%) | 0.38 |
| **Specificity n/N (95% CI)** | 1423/2325<br>61% (59-63%) | 143/321<br>45% (39-50%) | 1273/1987<br>64% (62-66%) | +19%<br>(14 to 25%) | <0.01 |

**Abbreviations:** CRP – C-reactive protein, MRS – microbiological reference standard, SXRS – sputum Xpert reference standard, CI – confidence interval, PWH – people living with HIV.

**Footnote**:
[a]19 participants had an unknown HIV status.
TB prevalence among PWH and people without HIV was 21% (p=0.90).

**Table S6. Diagnostic accuracy of CRP (5 mg/L cut-point) in reference to MRS and SXRS, by diabetes status.**

| | Overall | Diabetes status | | | |
|---|---|---|---|---|---|
| **In reference to MRS** | | **PWD** | **Non-diabetic** | **Difference** | **p-value** |
| **Sensitivity n/N (95% CI)** | 516/613 84% (81-87%) | 98/111 88% (81-94%) | 418/502 83% (80-86%) | -5% (-12 to 2%) | 0.19 |
| **Specificity n/N (95% CI)** | 1407/2291 61% (59-63%) | 158/271 58% (52-64%) | 1249/2020 62% (60-64%) | +4% (-3 to 10%) | 0.26 |
| **In reference to SXRS** | | | | | |
| **Sensitivity n/N (95% CI)** | 487/562 87% (84-89%) | 91/102 89% (82-95%) | 396/460 86% (83-89%) | -3% (-10 to 4%) | 0.40 |
| **Specificity n/N (95% CI)** | 1423/2325 61% (59-63%) | 159/277 57% (51-63%) | 1264/2048 62% (60-64%) | +5% (-2 to 11%) | 0.17 |

**Abbreviations:** CRP – C-reactive protein, MRS – microbiological reference standard,
SXRS – sputum Xpert reference standard, CI – confidence interval, PWD – people living with diabetes.

**Footnote**: TB prevalence among PWD was 29% vs. 20% among people without diabetes (p<0.01).

**Table S7. Diagnostic accuracy of CRP (5 mg/L cut-point) in reference to MRS and SXRS, by sex.**

| | Overall | Sex [a] | | | |
|---|---|---|---|---|---|
| **In reference to MRS** | | **Female** | **Male** | **Difference (95% CI)** | **p-value** |
| **Sensitivity n/N (95% CI)** | 516/613<br>84% (81-87%) | 144/183<br>79% (72-84%) | 372/430<br>87% (83-90%) | +8%<br>(1 to 15%) | 0.02 |
| **Specificity n/N (95% CI)** | 1407/2291<br>61% (59-63%) | 697/1125<br>62% (59-65%) | 709/1164<br>61% (58-64%) | -1%<br>(-5 to 3%) | 0.61 |
| **In reference to SXRS** | | | | | |
| **Sensitivity n/N (95% CI)** | 487/562<br>87% (84-89%) | 137/165<br>83% (76-88%) | 350/397<br>88% (85-91%) | +5%<br>(-1 to 12%) | 0.10 |
| **Specificity n/N (95% CI)** | 1423/2325<br>61% (59-63%) | 706/1138<br>62% (59-65%) | 716/1185<br>60% (58-63%) | -2%<br>(-6 to 2%) | 0.42 |

**Abbreviations:** CRP – C-reactive protein, MRS – microbiological reference standard, SXRS – sputum Xpert reference standard, CI – confidence interval, PWD – people living with diabetes.

**Footnote:**
[a]Two participants declined to state sex at birth.

**Table S8. Diagnostic accuracy of CRP (5 mg/L cut-point) in reference to MRS and SXRS, among diabetic sub-groups.**

| | **Sensitivity n/N (95% CI)** | | **Specificity n/N (95% CI)** | |
|---|---|---|---|---|
| | **In reference to MRS** | **In reference to SXRS** | **In reference to MRS** | **In reference to SXRS** |
| **All PWD** | 98/111<br>88% (81-94%) | 91/102<br>89% (82-95%) | 158/271<br>58% (52-64%) | 159/277<br>57% (51-63%) |
| **HbA1c ≥7.0%** | 52/63<br>83% (71-91%) | 49/58<br>85% (73-93%) | 71/125<br>57% (48-66%) | 72/128<br>56% (47-65%) |
| **HbA1c <7.0%** | 44/46<br>96% (85-100%) | 40/42<br>95% (84-99%) | 85/144<br>59% (51-67%) | 85/147<br>58% (49-66%) |
| **Previously diagnosed** | 51/61<br>84% (72-92%) | 48/56<br>86% (74-94%) | 115/172<br>67% (59-74%) | 117/176<br>67% (59-73%) |
| **Newly diagnosed[a]** | 47/50<br>94% (84-99%) | 43/46<br>94% (82-99%) | 43/99<br>43% (34-54%) | 42/101<br>42% (32-52%) |

**Abbreviations:** CRP – C-reactive protein, MRS – microbiological reference standard, SXRS – sputum Xpert reference standard, PWD – people with diabetes, HbA1c – hemoglobin A1c.

**Footnote:**
[a]Defined as HbA1c ≥6.5% and no prior reported history.

**Table S9. Univariate and adjusted ROC regressions for CRP in reference to MRS**.

| | Univariate | Multivariate | | | |
|---|---|---|---|---|---|
| | **Coefficient (95% CI)** | **Coefficient (95% CI)** | **SE** | **z** | **P>z** |
| **Female** | -0.093 (-0.27, 0.087) | -0.066 (-0.26, 0.13) | 0.10 | -0.68 | 0.50 |
| **Age** | -0.012 (-0.017, -0.006) | -0.0026 (-0.009, 0.004) | 0.0033 | -0.80 | 0.42 |
| **Uganda** | -- | -- | | | |
| **South Africa** | 0.066 (-0.19, 0.32) | 0.42 (0.13, 0.71) | 0.15 | 2.87 | 0.004 |
| **Philippines** | -0.66 (-0.96, -0.37) | -0.38 (-0.71, -0.040) | 0.17 | -2.19 | 0.028 |
| **Vietnam** | -0.71 (-0.93, -0.50) | -0.32 (-0.60, -0.047) | 0.14 | -2.30 | 0.022 |
| **India** | -0.48 (-0.81, -0.15) | 0.010 (-0.36, 0.38) | 0.19 | 0.05 | 0.96 |
| **Prior TB history** | -0.31 (-0.52, -0.093) | -0.22 (-0.45, 0.010) | 0.12 | -1.87 | 0.061 |
| **HIV-negative** | -- | -- | | | |
| **PWH CD4>200** | 0.022 (-0.29, 0.33) | -0.12 (-0.46, 0.22) | 0.17 | -0.70 | 0.49 |
| **PWH CD4≤200** | 0.49 (0.11, 0.86) | 0.43 (0.021, 0.84) | 0.21 | 2.06 | 0.039 |
| **BMI** | -0.061 (-0.086, -0.037) | -0.044 (-0.070, -0.018) | 0.013 | -3.32 | 0.001 |
| **No weight loss or weight loss ≤5kg** | -- | -- | | | |
| **Weight loss >5kg** | 0.41 (0.23, 0.58) | 0.049 (-0.16, 0.26) | 0.11 | 0.46 | 0.65 |
| **Heart rate** | 0.0098 (0.005, 0.015) | 0.0074 (0.002, 0.013) | 0.0027 | 2.76 | 0.006 |
| **Xpert-Negative/Very Low/Low** | -- | -- | | | |
| **Xpert-Medium/High** | 0.62 (0.45, 0.78) | 0.44 (0.27, 0.62) | 0.089 | 5.00 | <0.001 |
| *Constant (intercept)* | | *0.82 (0.019, 1.62)* | *0.41* | 2.01 | *0.045* |
| *Constant (slope)* | | *0.71 (0.66, 0.75)* | *0.023* | *30.87* | *<0.001* |

**Abbreviations:** ROC – receiver operating characteristic, CRP – C-reactive protein, MRS – microbiological reference standard, CI – confidence interval, SE – standard error, TB – tuberculosis, PWH – people living with HIV, BMI – body mass index.

**Footnote:** Variables that reflected a systemic inflammatory response were not included in the model (e.g. temperature, fever, night sweats).

**Table S10. Univariable and multivariable adjusted tobit regressions for elevated CRP on the logarithmic scale.**

| | Univariable (unadjusted) | Multivariable (adjusted for others in the model) | | |
|---|---|---|---|---|
| | Coefficient (95% CI) | Coefficient (95% CI) | SE | p-value |
| **Female** | -0.74 (-0.92, -0.57) | -0.40 (-0.55, -0.25) | 0.078 | <0.001 |
| **Age** | -0.015 (-0.020, -0.009) | -0.0016 (-0.007, -0.004) | -0.002 | 0.56 |
| **Uganda** | -- | -- | -- | -- |
| **South Africa** | -0.25 (-0.48, -0.015) | 0.85 (0.62, 1.08) | 0.12 | <0.001 |
| **Philippines** | -2.24 (-2.53, -1.95) | -1.22 (-1.49, -0.94) | 0.14 | <0.001 |
| **Vietnam** | -0.89 (-1.13, -0.65) | -0.57 (-0.81, -0.33) | 0.12 | <0.001 |
| **India** | -1.43 (-1.68, -1.17) | -0.70 (-0.95, -0.45) | 0.13 | <0.001 |
| **Prior TB history** | 0.24 (0.024, 0.45) | 0.080 (-0.099, 0.26) | 0.091 | 0.38 |
| **HIV-negative** | -- | -- | | |
| **PWH CD4>200** | 0.47 (0.19, 0.75) | -0.12 (-0.36, 0.13) | 0.13 | 0.35 |
| **PWH CD4≤200** | 1.88 (1.45, 2.30) | 0.54 (0.19, 0.88) | 0.18 | 0.002 |
| **Diabetes** | 0.31 (0.060, 0.57) | 0.53 (0.32, 0.75) | 0.11 | <0.001 |
| **BMI** | -0.12 (-0.14, -0.10) | -0.075 (-0.091, -0.059) | 0.008 | <0.001 |
| **No weight loss or weight loss ≤5kg** | -- | | | |
| **Weight loss >5kg** | 1.55 (1.36, 1.73) | 0.62 (0.44, 0.79) | 0.089 | <0.001 |
| **Heart rate** | 0.044 (0.039, 0.049) | 0.035 (0.030, 0.040) | 0.002 | <0.001 |
| **Smoked today** | -0.39 (-0.66, -0.12) | -0.61 (-0.85, -0.38) | 0.12 | <0.001 |

**Abbreviations:** CRP – C-reactive protein, CI – confidence interval, SE – standard error, TB – tuberculosis, PWH – people living with HIV, BMI – body mass index.
